# Association of Cognitive Deficits with Sociodemographic Characteristics among Adults with Post-COVID Conditions: Findings from the United States Household Pulse Survey

**DOI:** 10.1101/2023.09.22.23295981

**Authors:** Daniel J Wu, Nianjun Liu

**Author notes:** Corresponding authors: Daniel J. Wu, Shadow Creek High School, 11850 Broadway St., Pearland, Texas 77584., Nianjun Liu, Ph.D. Department of Epidemiology and Biostatistics, Indiana University School of Public Health, 1025 E. Seventh Street, Suite 111, Bloomington, IN 47405-7109.

## Abstract

**Background:** People infected with COVID-19 may continue to experience symptoms for several weeks or even months after acute infection, a condition known as long COVID. Cognitive problems such as memory loss are among the most commonly reported symptoms of long COVID. However, a comprehensive evaluation on the risks of cognitive decline following COVID infection among different sociodemographic groups has not been undertaken at the national level in the United States.

**Methods:** We conducted a secondary analysis on the datasets from U.S. Census Bureau Household Pulse Survey, encompassing the data collected from June 1, 2022 to December 19, 2022. Based on a cohort of 385,370 individuals aged 18 or older, we employed logistic regression analyses to examine the association between self-reported cognitive deficits and different sociodemographic factors among individuals with long COVID conditions.

**Results:** Among individuals aged 18 or older, 44.7% percent of survey respondents report having been diagnosed with COVID in the past, and 29.0% of those with previous COVID infection experienced long COVID symptoms lasting for more than 3 months. We have demonstrated that individuals with long COVID had significantly higher risk of experiencing cognitive deficits compared to those with no history of COVID infection. Furthermore, females, young adults, people with multiple races, or low levels of education attainment are at high risk of cognitive deficits if they experience long COVID. At the state level, the prevalence of cognitive deficits among long COVID patients varied across different US states, with the highest prevalence in West Virginia and Kentucky, and the lowest prevalence in Connecticut and Maryland. The variation could be due to differences in racial composition and education level among long COVID patients in the four states.

**Conclusions:** The risks of cognitive deficits among adults with post-COVID conditions are substantial. Various sociodemographic groups can have different risks of developing cognitive deficits after experiencing long COVID. Findings of this large-scale study can help identify sociodemographic groups at higher risk of cognitive deficits, and facilitate medical interventions and guide resource allocation to target populations at risk and to prioritize areas with a high rate of cognitive decline.

## Introduction

COVID-19 caused by severe acute respiratory syndrome coronavirus 2 (SARS-CoV-2) has posed a major challenge to public health worldwide. As of July 2023, there have been more than 760 million confirmed cases of COVID-19, including 6.9 million deaths worldwide [1]. While recent unprecedented progress in clinical research and vaccine development has significantly helped the prevention of COVID-19, accumulating evidence suggests that COVID survivors experience persistent symptoms after initial infection [2–4]. Many survivors are often diagnosed with post-acute sequelae of COVID-19, or simply long COVID, broadly defined as persistent symptoms and clinical abnormalities lasting beyond the first 30 days of infection [5, 6]. Long COVID can affect multiple organ systems, including heart, brain, kidney, and gastrointestinal tract [3, 7–10]. In 2021, long COVID was recognized as a condition that could result in a disability under the Americans with Disabilities Act [11]. While current studies consistently demonstrate that the severity of acute symptoms is a significant risk factor for long COVID, they have yielded controversial results on how the risk varies among different demographic groups. For example, Hastie et al. claimed that the risk of long COVID was higher in females, older adults, and white ethnicity [4]. In contrast, Wu et al. found no significant association between long COVID and sociodemographic factors [12]. These controversial findings could be due to the fact that long COVID reflects a wide spectrum of symptoms, and each symptom may manifest differently in various demographic groups. For example, symptoms such as sleep disorders and headache appear more pronounced in younger adults, while others, such as anxiety and dyspnea, are more prevalent in adults older than 70 [13]. Because long COVID manifests as a heterogeneous problem, it would be better to focus on specific symptoms when studying their associated risk factors. Our study will focus on cognitive symptoms characterized as loss of memory and impaired concentration, one of the most common and distressing features of long COVID conditions [14].

Recent neuroimaging studies have demonstrated that COVID infection may cause widespread structural abnormalities in the brain, which may help explain the lasting cognitive dysfunction among people with long COVID conditions [15, 16]. The relationship between cognitive impairment and COVID care sites was assessed within a cohort of 740 individuals aged 18 and above, all of whom had contracted COVID-19 [17]. According to the study, seven months after their COVID-19 infection, 15% of patients who had been hospitalized, 10% of those requiring Emergency Department admission, and 8% of outpatients exhibited signs of impaired working memory [17]. A large study involving around 81,000 individuals, primarily from the United Kingdom, has reported consistent findings concerning the influence of COVID infection on cognitive function. The study indicates an elevated risk of cognitive decline that correlates with the severity of acute COVID infection. Notably, patients who required hospitalization and ventilator support displayed the most significant cognitive regression [18]. A recent study employed the US Department of Veterans Affairs healthcare databases to estimate risks and burdens of neurological disorders at 12 months following initial COVID infection. It was demonstrated that individuals experiencing ongoing long COVID symptoms after the initial onset performed significantly worse than those who had fully recovered, suggesting a common occurrence of memory problems among individuals with long COVID conditions [19].

Overall, the existing academic literature has demonstrated that long COVID patients exhibit neurocognitive impairments, including memory deficits, potentially due to brain structural and functional changes after COVID infection. However, most studies utilized European cohorts. While there are a few studies utilizing US cohorts, they typically have only incorporated relatively small sample sizes or limited diversity in demographic groups [17, 18, 20]. A comprehensive evaluation on the risks of cognitive problems among individuals with long COVID condition has not been undertaken at the national level in the United States. Beginning in June 2022, the National Center for Health Statistics collaborated with the United States Census Bureau to include long COVID-related questions in the experimental Household Pulse Survey [21]. This online survey makes the first large-scale dataset about the prevalence of long COVID, providing a unique opportunity to study the impacts of long COVID on cognitive functions among the US population at the nationwide level. In this study, by leveraging the responses from this national survey, we assess the prevalence of cognitive deficits among adult COVID survivors in the United States and compare the risks of severe cognitive deficits across different sociodemographic groups.

## Methods

### Data Source

The Household Pulse Survey (HPS) was initially launched in April 2020 and aims to quickly collect real-time data to explore the impacts of COVID-19 on American households and individuals [22]. It is a 20-minute online survey randomly selecting people aged 18 years or older to participate. Unique phone numbers and email addresses are assigned to only one household in a de-duplication process. Households were randomly sampled from the US Census Bureau’s Master Address File and received the survey via text or email. As of August 2023, the HPS continues with a two-weeks on, two-weeks off collection and distribution approach. The survey asks questions related to physical and mental health, transportation, childcare, as well as detailed demographic information of individuals. Starting from June 2022, a set of questions to assess the prevalence of long COVID were added to the survey [21]. The questions asked include “Did you have any symptoms lasting 3 months or longer that you did not have prior to having coronavirus or COVID-19?” While the survey yielded low response rate of about 6%, its inclusion of long COVID related questions and a number of sociodemographic variables allows us to study the association between peoples’ cognitive status and their sociodemographic characteristics among people with long COVID conditions at the US population level.

### Data Collection

We downloaded seven microdata files from HPS data collection phases 3.5 to 3.7, spanning the timeframe between June 1, 2022 and December 19, 2022 [23]. A consolidated dataset was generated by merging these microdata files. The dataset included survey responses from 392,073 individuals. After excluding the individuals with missing responses to the COVID-related questions, we obtained a cohort of 385,370 individuals. With regards to the cognitive outcome, we examined the survey responses to the question in the cognition domain: “Do you have difficulty remembering or concentrating? (1) No - no difficulty; (2) Yes - some difficulty; (3) Yes - a lot of difficulty; and, (4) Cannot do at all.” This question was recommended by the United Nations to identify disability in national censuses [24]. Following the United Nations’ guidelines, current literature identify people who answer “a lot of difficulty” or “cannot do at all” as having the disability (eg. [25, 26]). In this study, we categorize the cognitive outcome into two levels for further analysis. Individuals who answer “no difficulty” or “some difficulty” are categorized as not having severe cognitive deficits, while individuals who answer “a lot of difficulty” or “cannot do it at all” are classified as having severe cognitive deficits.

We also retrieved self-reported survey responses regarding individuals’ sociodemographic status, including gender at birth, race/ethnicity, age (based on the year of birth), educational attainment, region and state of residence, to determine their association with the self-reported cognitive deficits among individuals. It was demonstrated that these sociodemographic factors were associated with long COVID conditions based on a large-scale study involving the Scottish population [4]. In light of this, we chose to incorporate these variables into this study to investigate their potential association with cognitive deficit within the US population.

### Data Analysis

All data analyses and visualization were performed using R, version 4.2.2 [27]. All p-values were two-sided and p-values less than 0.05 were considered statistically significant. First, for assessing the long COVID prevalence, chi-square tests were used to investigate group differences in relation to long COVID, categorized by various sociodemographic variables including gender, age, race/ethnicity, region of residence, and educational attainment level.

We then used a binary logistic regression model to quantify the association between the dichotomous outcome of cognitive deficit and long COVID status. Both univariate logistic regression model and the adjusted model to account for other potential covariates (i.e., gender, age, race/ethnicity, education level, and regions) were run. We also used multivariable logistic regression model to further assess various sociodemographic factors associated with cognitive deficit among long COVID patients. The odds ratio for each sociodemographic factor quantifies the strength of the association.

Lastly, we compared the prevalence of self-reported cognitive deficits among long COVID patients across all 50 US states and the District of Columbia. We further analyzed the sociodemographic characteristics of two states with the highest prevalence and two states with the lowest prevalence of cognitive deficits. Chi-squared tests were used to determine whether there were significant differences in the sociodemographic distributions among these states.

## Results

### Characteristics of Survey Participants

Among the 385,370 participants in our dataset, 212,992 (55.3%) reported no previous COVID infection. Out of the remaining 172,378 individuals who reported previous COVID infection, 50,038 participants experienced long COVID symptoms lasting for more than 3 months, comprising 29.0% of the infected group and 13.0% of all survey participants (Table 1). Five categorical variables, namely gender at birth, race/ethnicity, age, region of residence, and educational attainment, were derived from the response categories in the survey. We then disaggregated the individuals into their respective categories and conducted chi-squared tests to investigate group differences in relation to long COVID status. The results demonstrate that long COVID disproportionally affects various sociodemographic groups. For example, among participants reporting long COVID, 69.1% are females, while among all survey participants, only 56.9% are females. The difference suggests that females are more likely to experience long COVID than males (p < 0.0001). Analysis on race/ethnicity suggests that long COVID is most prevalent among Hispanics (12.0%) and least prevalent among non-Hispanic Asian (2.8%). For age distribution, we created four major age groups and classified the participants into these groups: young adults (18-34), early middle-aged (35-49), late middle-aged (50-64), and old adults (65+). These age cut-off points were chosen according to the general developmental stage in adult life span [28, 29]. Our estimation indicates that just 15.9% of individuals who reported long COVID symptoms are 65 years and older. This represents the lowest prevalence of long COVID among all four age groups (p < 0.0001). In addition, long COVID is more common among individuals without a college degree (55.3%), compared to those with a graduate degree (19.2%). Lastly, only 13.3% of long COVID patients reside in the Northeast region, significantly lower than the other three regions of the US (p< 0.0001).

**Table 1.**
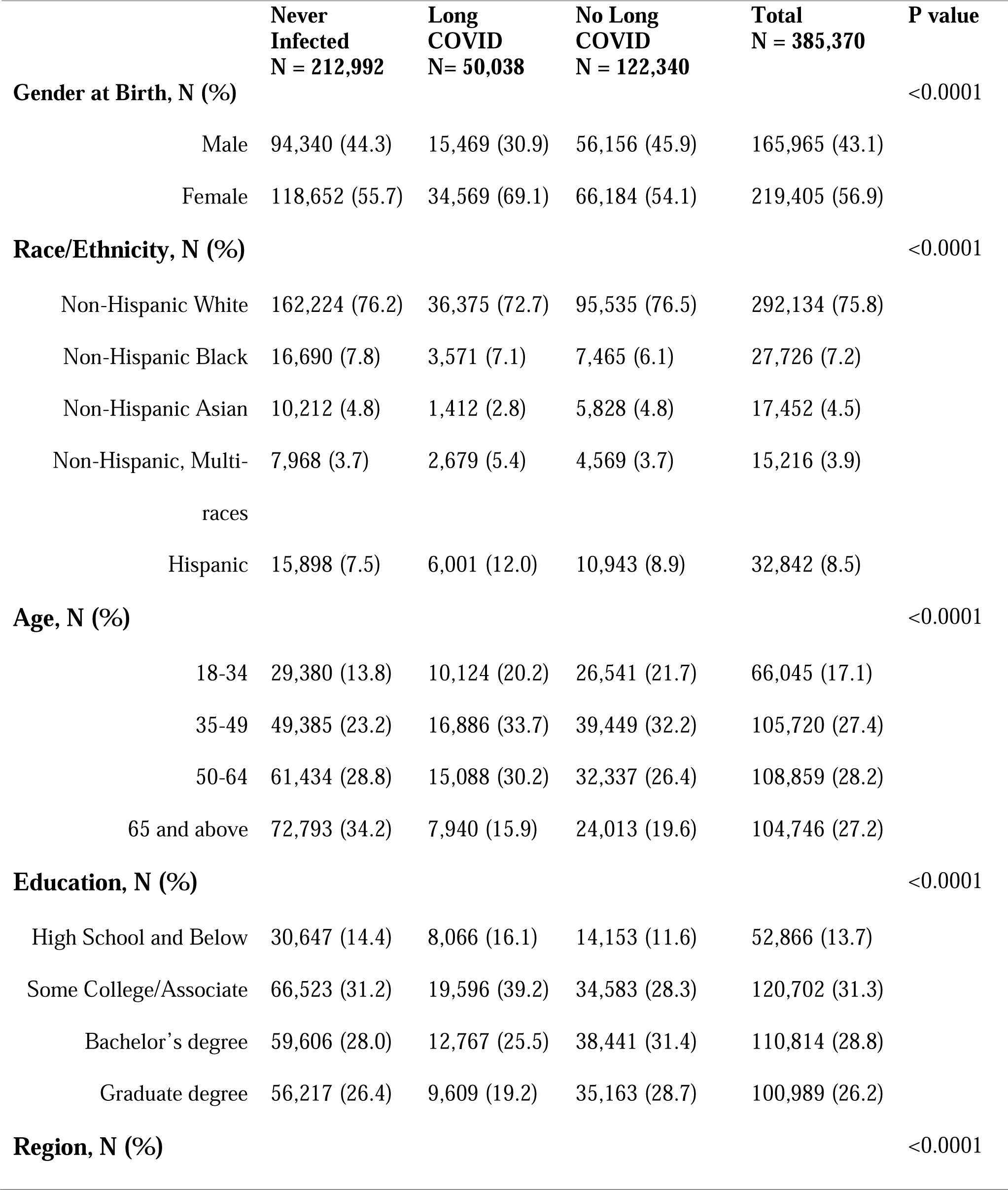
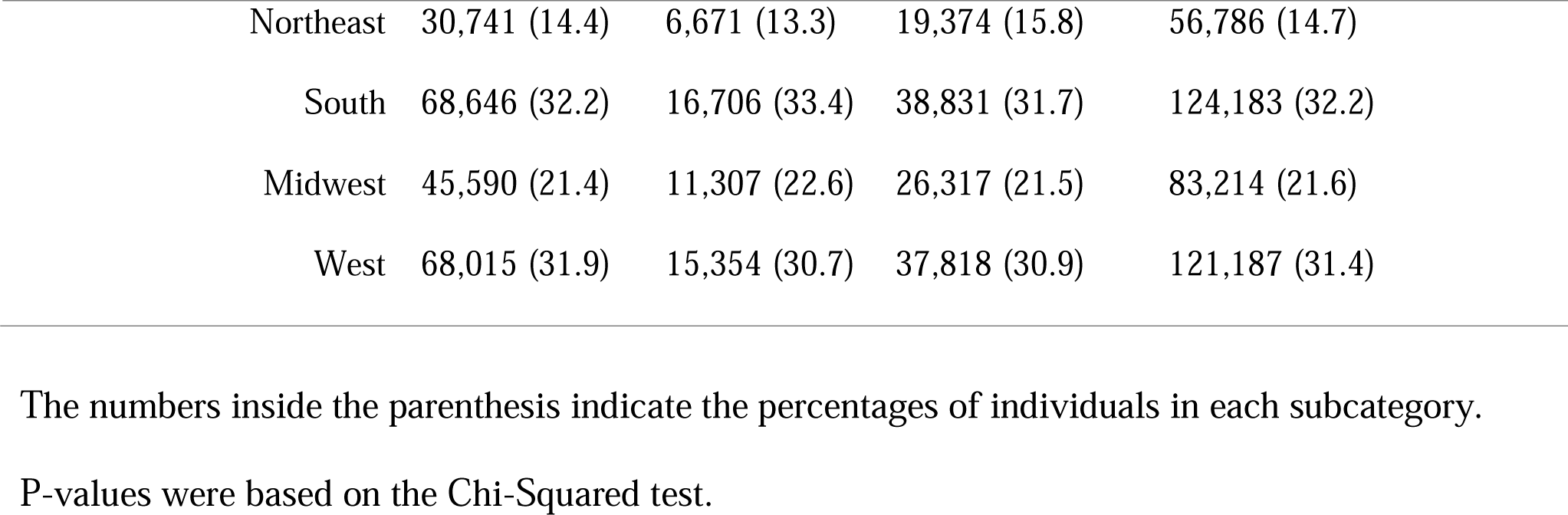
Summarized sociodemographic characteristics of survey participants.

|  |  | **Never**<br>**Infected**<br>**N = 212,992** | **Long**<br>**COVID**<br>**N= 50,038** | **No Long**<br>**COVID**<br>**N = 122,340** | **Total**<br>**N = 385,370** | **P value** |
| --- | --- | --- | --- | --- | --- | --- |
| **Gender at Birth, N (%)** |  |  |  |  |  | <0.0001 |
|  | Male | 94,340 (44.3) | 15,469 (30.9) | 56,156 (45.9) | 165,965 (43.1) |  |
|  | Female | 118,652 (55.7) | 34,569 (69.1) | 66,184 (54.1) | 219,405 (56.9) |  |
| **Race/Ethnicity, N (%)** |  |  |  |  |  | <0.0001 |
|  | Non-Hispanic White | 162,224 (76.2) | 36,375 (72.7) | 95,535 (76.5) | 292,134 (75.8) |  |
|  | Non-Hispanic Black | 16,690 (7.8) | 3,571 (7.1) | 7,465 (6.1) | 27,726 (7.2) |  |
|  | Non-Hispanic Asian | 10,212 (4.8) | 1,412 (2.8) | 5,828 (4.8) | 17,452 (4.5) |  |
|  | Non-Hispanic, Multi-races | 7,968 (3.7) | 2,679 (5.4) | 4,569 (3.7) | 15,216 (3.9) |  |
|  | Hispanic | 15,898 (7.5) | 6,001 (12.0) | 10,943 (8.9) | 32,842 (8.5) |  |
| **Age, N (%)** |  |  |  |  |  | <0.0001 |
|  | 18-34 | 29,380 (13.8) | 10,124 (20.2) | 26,541 (21.7) | 66,045 (17.1) |  |
|  | 35-49 | 49,385 (23.2) | 16,886 (33.7) | 39,449 (32.2) | 105,720 (27.4) |  |
|  | 50-64 | 61,434 (28.8) | 15,088 (30.2) | 32,337 (26.4) | 108,859 (28.2) |  |
|  | 65 and above | 72,793 (34.2) | 7,940 (15.9) | 24,013 (19.6) | 104,746 (27.2) |  |
| **Education, N (%)** |  |  |  |  |  | <0.0001 |
|  | High School and Below | 30,647 (14.4) | 8,066 (16.1) | 14,153 (11.6) | 52,866 (13.7) |  |
|  | Some College/Associate | 66,523 (31.2) | 19,596 (39.2) | 34,583 (28.3) | 120,702 (31.3) |  |
|  | Bachelor's degree | 59,606 (28.0) | 12,767 (25.5) | 38,441 (31.4) | 110,814 (28.8) |  |
|  | Graduate degree | 56,217 (26.4) | 9,609 (19.2) | 35,163 (28.7) | 100,989 (26.2) |  |
| **Region, N (%)** |  |  |  |  |  | <0.0001 |

|  |  |  |  |  |
| --- | --- | --- | --- | --- |
| Northeast | 30,741 (14.4) | 6,671 (13.3) | 19,374 (15.8) | 56,786 (14.7) |
| South | 68,646 (32.2) | 16,706 (33.4) | 38,831 (31.7) | 124,183 (32.2) |
| Midwest | 45,590 (21.4) | 11,307 (22.6) | 26,317 (21.5) | 83,214 (21.6) |
| West | 68,015 (31.9) | 15,354 (30.7) | 37,818 (30.9) | 121,187 (31.4) |

The numbers inside the parenthesis indicate the percentages of individuals in each subcategory.
P-values were based on the Chi-Squared test.

### Association between Cognitive Deficits and Long COVID

We evaluated the association between cognitive deficit and the long COVID status (Table 2). After excluding nonrespondents on the cognitive outcome question, we obtained a cohort of 335,395 individuals. Compared to individuals who never had COVID, those who have experienced long COVID have a significantly higher odds of experiencing severe cognitive deficit, as indicated by the univariate model (OR=3.37, 95% CI 3.26-3.49). Even after accounting for sociodemographic variables, the long COVID group still shows higher risk of cognitive deficit compared to the group who had never been infected (adjusted OR=2.87, 95% CI 2.78-2.97). However, people who had previous COVID infection but did not report long COVID symptoms only have a slightly increased risk of cognitive deficit (adjusted OR = 1.08, 95% CI 1.05-1.12).). We further examined the association between the severity of COVID acute infection and the odds of reporting severe cognitive deficit. While asymptomatic symptoms were not associated with an increased odds of cognitive deficit (OR = 1.01, p-value = 0.75), people who had mild, moderate or severe symptoms during the acute infection stage were significantly more likely to report cognitive disability (Table S1). The significance of the association increases with the severity of symptoms. Compared to those who had never been infected with COVID, people with mild, moderate, or severe symptoms had 1.24, 1.32, and 3.07 times the odds of having cognitive deficit (all p values are smaller than 0.0001), respectively (Table S1).

**Table 2.** Logistic regression analysis of associations between cognitive deficit and COVID infection.

|  | Severe Cognitive Deficits, N |  | Univariate |  | Adjusted |  |
| --- | --- | --- | --- | --- | --- | --- |
|  | Yes | No | OR (95% CI) | p-value | OR (95% CI) | p-value |
| **Never Infected** | 9,415 | 175,436 | 1.0 | Reference | 1.0 | Reference |
| **No Long COVID** | 5,998 | 100,933 | 1.11 (1.07, 1.14) | <0.0001 | 1.08 (1.05, 1.12) | <0.0001 |
| **Long COVID** | 6,683 | 36,930 | 3.37 (3.26, 3.49) | <0.0001 | 2.87 (2.78, 2.97) | <0.0001 |

N, the number of people. OR, odds ratio. CI, confidence interval. Adjusted, model controlled for gender, race/ethnicity, age, education, and region. Odds ratio calculation is referent to people who never had COVID.

### Association between Cognitive Deficit and Sociodemographic Factors

The risks of cognitive deficit are different among various sociodemographic groups who have reported long COVID symptoms (Figure 1 and Table S2). Compared to males, females with long COVID symptoms have 1.32 higher odds to experience severe cognitive deficit (OR= 1.32, p-value < 0.0001). There is no significant difference in the risks of cognitive deficit between non-Hispanic whites and Hispanics (p-value = 0.467). However, compared to non-Hispanic whites, individuals with multi-races are more likely to report severe cognitive deficits (OR = 1.27, p-value < 0.0001), while non-Hispanic Asians (OR = 0.65, p-value < 0.0001) and Blacks (OR = 0.85, p-value = 0.003) are less likely. Among the four age groups observed, young adults aged 18-34 have the highest odds of reporting severe cognitive deficit, while adults aged 50-64 have the lowest odds (OR = 0.65, p-value < 0.0001). Additionally, individuals with a Bachelor’s (OR = 0.72, p-value < 0.0001) or a graduate degree (OR = 0.63, p-value < 0.0001) have significantly lower odds of cognitive deficit than those with a high school diploma or less. Finally, although no significant differences were observed between the Northeast, West, and Midwest, individuals living in the South region have higher odds of cognitive deficits than those living in the other three regions (OR = 1.21, p-value <0.0001).

**Figure 1.**
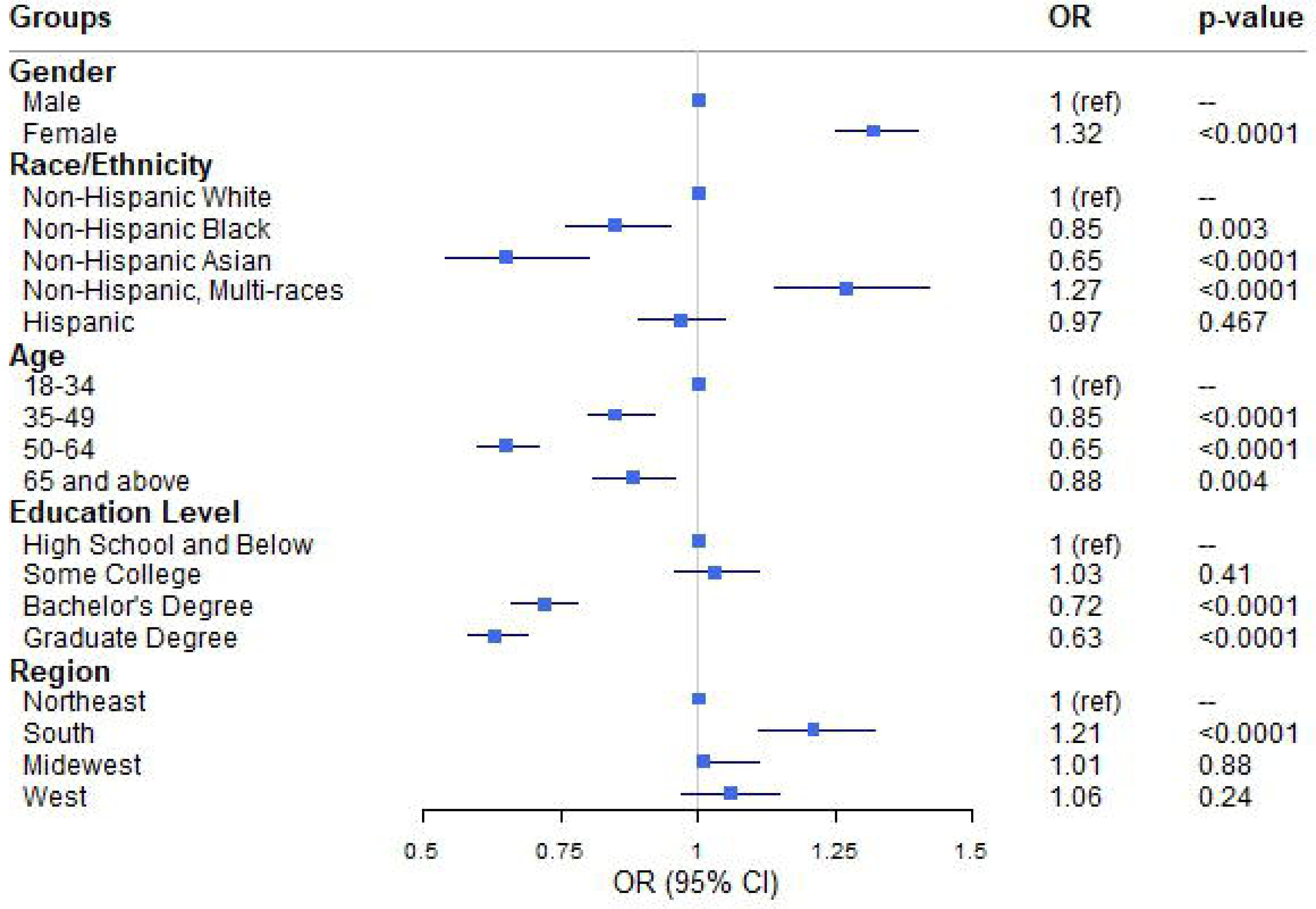
Forest plot representing odds ratio of different sociodemographic variables. Odds ratios are calculated with adjusted logistic regression analyses that are controlled for the other categorical variables in the plot.

### Prevalence of Cognitive Deficit in Different States

We further examined whether there were differences in the prevalence of cognitive deficit among long COVID patients across various US states. We calculated the percentages of long COVID patients reporting severe cognitive deficits in each state and used a color-shaded US map to visualize the results (Figure 2). It is evident that Kentucky (19.6%) and West Virginia (20.0%) had the highest rates of cognitive deficits among individuals with long COVID symptoms. In contrast, Connecticut (11.6%) and Maryland (12.4%) exhibited the lowest percentages (Figure 2, Table S3).

**Figure 2.**
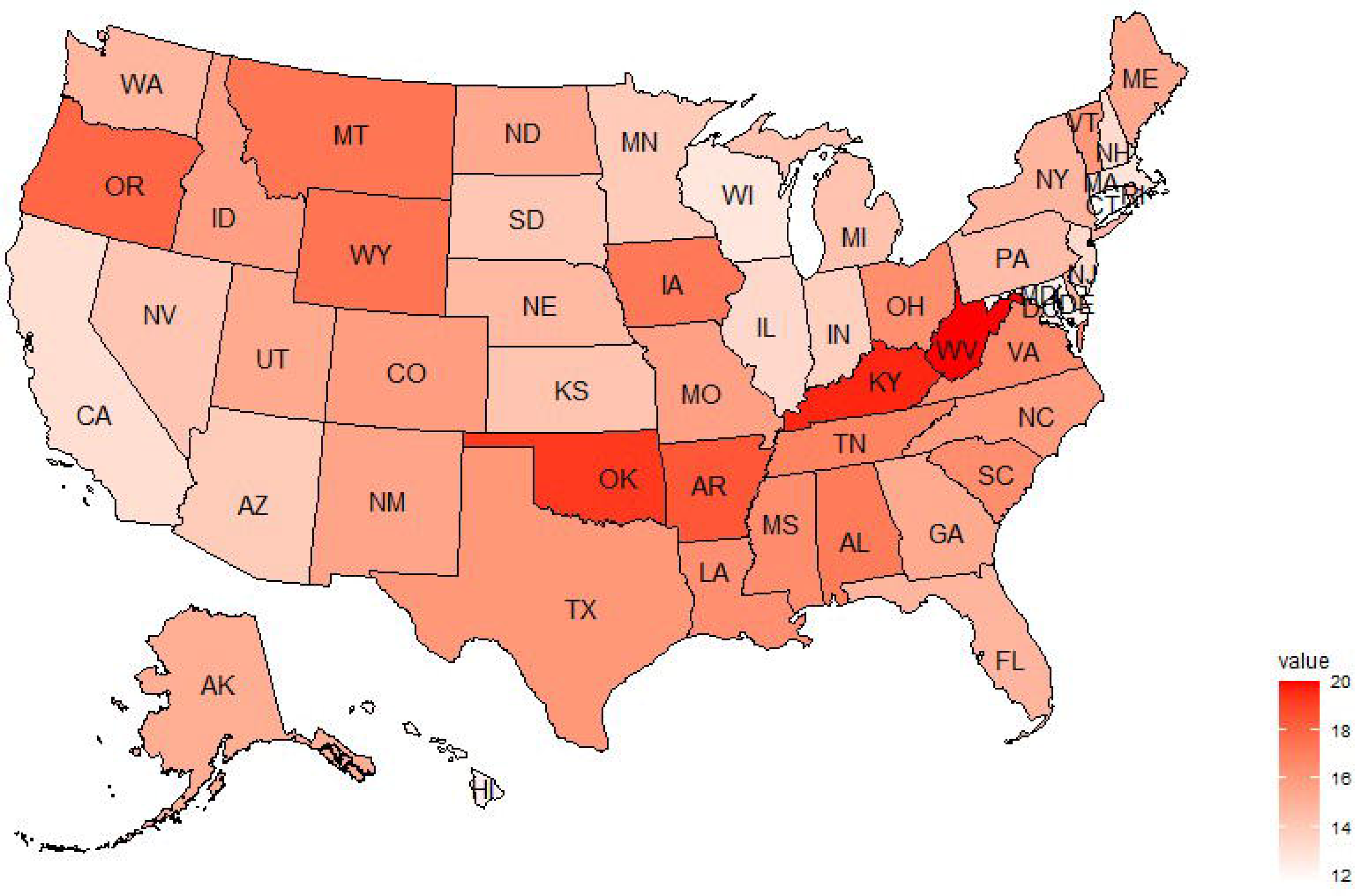
Prevalence of cognitive deficit reported by long COVID patients in various US states. The shading of each state in the US map corresponds to the percentage of long COVID patients reporting severe cognitive deficits in the state.

To investigate potential risk factors contributing to the state variations, we compared the sociodemographic characteristics of long COVID patients in the four states (CT, MD, KY, and WV). It indicated there were no significant differences in the gender (p-value = 0.89) or age distribution (p-value = 0.23) among the long COVID patients across these four states (Figure 3, Table S4). However, the race distribution among long COVID patients is significantly different in the four states (chi-squared = 415.9, p < 0.0001). According to Table S4, the proportion of non-Hispanic whites among long COVID patients in Connecticut (70.7%) and Maryland (59.6%) is much lower compared to Kentucky (89.6%) and West Virginia (92.5%). In contrast, Connecticut and Maryland have a higher percentage of non-Hispanic Asians and Blacks compared to Kentucky and West Virginia (Figure 4A, Table S4). In addition, There are significant state differences in the proportion of people with low education among the four states (chi-squared = 80.4, p < 0.0001). Figure 4B indicates that people tend to have high education levels in Connecticut and Maryland. Only 51.5% and 42.1% of long COVID patients have no Bachelor degree in Connecticut and Maryland, respectively, much lower than the percentages in Kentucky (59.9%) and West Virginia (63.7%) (Table S4). These results further support the previous finding that non-Hispanic whites are more likely to report cognitive deficits compared to Blacks and Asians, and individuals with low educational attainment have higher risk of experiencing cognitive deficits among long COVID patients.

**Figure 3.**
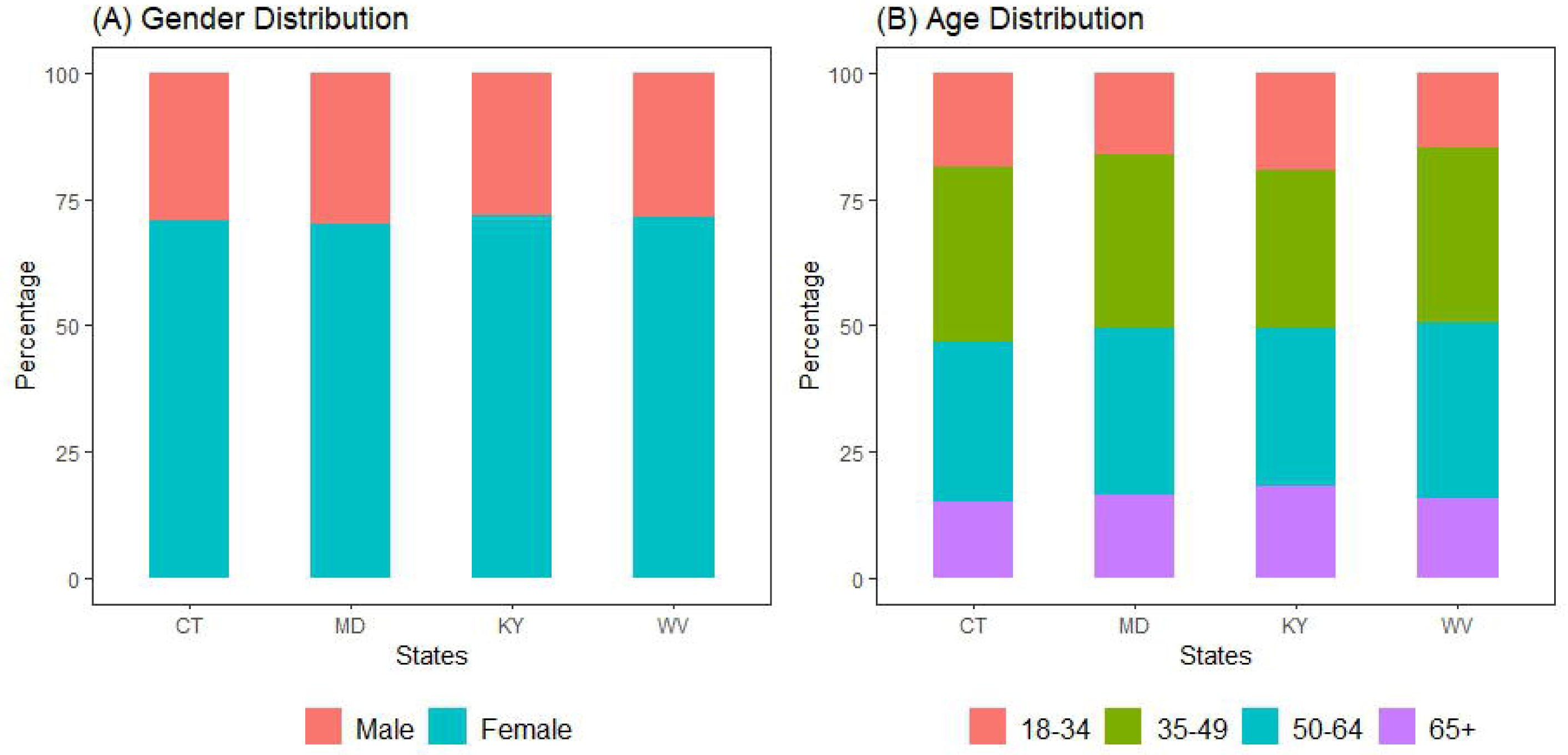
Gender and age distribution among long COVID patients in four states.

**Figure 4.**
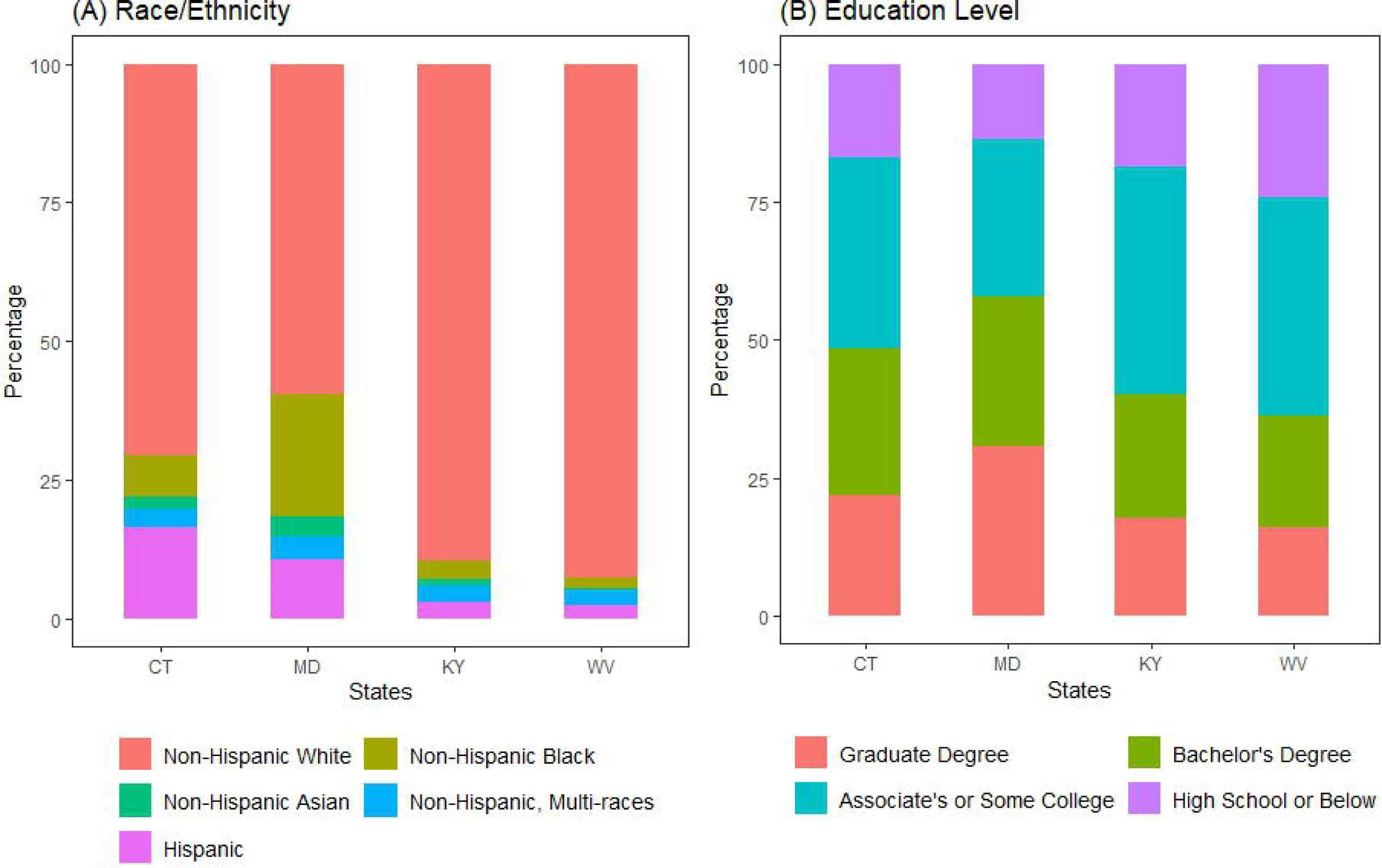
Racial composition and educational attainment levels among long COVID patients in four states.

## Discussion

In the analysis of 385, 370 survey respondents between June and December of 2022, we estimated that approximately 29% of individuals who had previously contracted COVID also experienced long COVID symptoms lasting more than three months. The estimation of long COVID prevalence in the US national level is lower than that reported by a Scottish study, where more than 42% of COVID survivors experienced persistent symptoms after six months [4]. It is important to highlight that the questionnaire utilized in the Scottish study included a total of 33 symptoms, all classified as long COVID conditions. In contrast, the House Pulse Survey our study relied on includes only 12 symptoms. Therefore, the difference in how long COVID conditions are defined and the range of persistent symptoms between the Scottish study and ours could potentially contribute to the divergent results. Our results also demonstrate the prevalence of long COVID tend to be low in the oldest age group (65+), consistent with a recent study based on a population-representative sample of 3042 US adults [30]. The lower prevalence of long COVID among older people can be due to several factors such as survivor bias, lower rates of virus exposure, and higher rates of vaccination of old adults [31].

Our study suggests that people with long COVID are at increased risk of cognitive deficits in memory and concentration and the risk correlates with the severity of COVID symptoms during acute infection phase. More specifically, for asymptomatic patients, they do not have an increased risk compared to those who never got infected. These findings corroborate with the current existing studies investigating the impact of COVID on cognitive status [32]. Furthermore, we have examined the association between different sociodemographic factors and cognitive deficits in long COVID patients and found that females had higher risk of experiencing cognitive deficit than males. However, in a recent study of 72 mild-to-moderate COVID survivors, Henneghan et al. found no gender difference in the overall cognitive function measured by a neuropsychological test [33]. We note that their study only included a small sample size and focuses on mild-to-moderate cases, therefore, potential gender effects in cognitive outcomes may not be detected. In addition, our results suggest that the risk of cognitive deficit is the highest in the age group of 18-34 years compared to other age groups. A large cohort study based on patients in the US of Veterans Affairs healthcare database reached a similar conclusion, demonstrating that the risks of cognition and memory disorders decreased as age increased [19]. Given that young adulthood is still in the stage of cognitive development, even a mild disruption of underlying biological processes caused by COVID can have significant consequences on cognitive outcomes, possibly providing an explanation for the strong association between memory deficit and young age [34].

Among different race groups, this study show that the risk of cognitive deficit is the highest among people with multiple races and the lowest among Asians. Some prior studies on the association of long COVID with memory disorders did not include subjects with diverse races/ethnicity, potentially limiting the scope of their findings [35, 36]. There may be multiple factors for the racial disparity on cognitive outcome shown in this study. One possibility is that the biological effects of COVID-19 may differ due to genetic differences in various racial groups, consistent with a review on the genetic insight for COVID [37]. Furthermore, we suggest that individuals with the lowest education level (high school diploma or below) have a significantly higher risk of developing cognitive deficits compared to other subgroups. Individuals with lower levels of education often have lower economic status [38]. Their economic disadvantages limit their access to healthcare and making them less likely to receive timely and effective treatment for COVID-19, potentially increasing their risk for developing cognitive symptoms. Alternatively, the result about the impact of educational level could be explained by the passive cognitive reserve theory [39]. According to this theory, people with higher levels of education tend to have greater cognitive reserve, which can help maintain their cognitive performance level for longer periods of time and reduce the risk of cognitive decline and dementia [40, 41].

As our research represents the first major effort to investigate the prevalence of cognitive deficit among long COVID patients across different US states, it is difficult to find other similar researches to compare with. However, our results corroborate other findings in this study. Since race and educational levels are risk factors associated with cognitive outcome, the prevalence of cognitive deficits among US states can differ significantly if there are variations in race distribution and education levels among these states. Therefore, this finding not only corroborates that individuals’ sociodemographic characteristics contribute to their cognitive outcomes, but also suggests regional variability among different US states.

COVID-19 pandemic remains a constantly evolving situation. As more treatment for acute COVID becomes available and the number of individuals receiving vaccines and boosters increases, the epidemiology of long COVID may change over time [42]. Therefore, it is important to conduct further research to evaluate the potential impact of vaccination on reducing the risk of cognitive deficits among patients who have breakthrough infection, as well as to identify which sociodemographic groups may benefit the most from vaccination. This information can help guide public health planning and vaccine distribution efforts. Furthermore, while the scope of this study focuses on individuals aged 18 and above, future research can be conducted to study cognitive decline among children with long COVID as well. Cognitive deficits can have a significant impact on children’ academic and social functioning, potentially affecting their future opportunities. Understanding the extent of cognitive decline in this population group will help identify risk factors and guide the development of interventions, such as cognitive rehabilitation, to improve outcomes for affected children. Therefore, it is important to conduct research on the long term effects of COVID in all age groups to address the broader impact of the disease.

### Limitations

This study has several limitations. First, the study relies on self-reported survey data, including the response to questions of long COVID symptoms and memory deficits. Compared to the objective physiological or cognitive tests, the self-reported responses can be biased due to personal interpretation of the questions. As a result, the accuracy of the assessment on the association between the risk of cognitive deficit and long COVID across various sociodemgraphic groups might be affected by the response bias. Second, the Household Pulse Survey has a relatively low response rate of about 6% and the survey data may be subject to sampling bias. For example, individuals who have more severe symptoms may be more likely to participate in this survey, leading to an overrepresentation of more severe cases in the data. Finally, although the study demonstrates strong associations between individuals’ long COVID status and their cognitive outcome, we cannot definitely establish a causal relationship between long COVID and cognitive deficit. Other confounding variables, such as individuals’ pre-existing health conditions, may also play a role in their cognitive outcome. Not considering pre-existing symptoms prior to COVID infection could result in an overestimation of the impact of long COVID. Therefore, it is important to supplement this study with other types of research, such as longitudinal studies, to better understand the impacts of long COVID on cognitive functions.

### Conclusions

Long COVID has become a world-wide health issue. This study based on a large cohort representative of the US adult population has found significant differences in the cognitive outcomes associated with COVID infections among various sociodemographic groups. Females, young adults, individuals with multiple races, or low levels of education attainment are at high risk of cognitive deficits if they experience post-COVID conditions, highlighting the importance of considering both biological and non-biological factors in understanding differential cognitive outcomes of COVID infection. The difference in the prevalence of cognitive deficit among different US states also prompt further inquiry on specific regions. Such studies may guide future diagnosis, interventions and treatments to improve cognitive outcomes among people with post-COVID conditions.

## Data Availability

All data produced in the present work are contained in the manuscript.

https://www.census.gov/programs-surveys/household-pulse-survey/datasets.html

## List of abbreviations

COVID-19: Coronavirus Disease 2019
HPS: Household Pulse Survey
WHO: World Health Organization
OR: odds ratio
CI: Confidence interval

## Declarations

### Ethics approval and consent to participate

Data for the study were obtained from a national survey conducted by the United States Census Bureau. Ethics approval was waived because the data are de-identified and publicly available. The study was a secondary analysis of the survey data and therefore consent to participate is not applicable.

## Consent for publication

### Availability of data and materials

The microdata files containing individual responses to the Household Pulse Survey are available for download from https://www.census.gov/programs-surveys/household-pulse-survey/datasets.html

## Competing interests

### Author’s Contributions

DJW and NL designed the research. DJW performed data collection and analyses. NL provided technical and material support, as well as supervision and mentorship. DJW and NL interpreted results and wrote the manuscript together. All authors read and approved the final manuscript.

## Acknowledgements

We would like to thank Pamela Blades for her valuable guidance throughout the study.

