## Additional File for "Association of Cognitive Deficits with Sociodemographic Characteristics among Adults with Post-COVID Conditions: Findings from the United States Household Pulse Survey"

**Table S1.** Logistic regression analysis of associations between severe cognition deficit and acute COVID infection severity, referent to people never infected.

| | Univariate model | | Adjusted model | |
|---|---|---|---|---|
| | **OR (95% CI)** | **p-value** | **OR (95% CI)** | **p-value** |
| **Asymptomatic** | 1.12 (0.99, 1.26) | 0.05 | 1.01 (0.91, 1.14) | 0.75 |
| **Mild** | 1.46 (1.40, 1.52) | <0.0001 | 1.24 (1.19, 1.30) | <0.0001 |
| **Moderate** | 1.49 (1.43, 1.55) | <0.0001 | 1.32 (1.26, 1.37) | <0.0001 |
| **Severe** | 3.97 (3.78, 4.17) | <0.0001 | 3.07 (2.91, 3.22) | <0.0001 |

OR: odds ratio. Adjusted: model adjusted for age, gender, race/ethnicity, education, and region.

**Table S2.** Association between cognitive deficit and sociodemographic factors among long COVID patients

| | **Severe Cognitive Deficit** | | **OR (95% CI)** | **p-value** |
|---|---|---|---|---|
| | **Yes** | **No** | | |
| **Gender at Birth, N** | | | | |
| Male | 1,738 | 11,857 | 1.0 | Reference |
| Female | 4,945 | 25,073 | 1.32 (1.25, 1.40) | <0.0001 |
| **Race/Ethnicity, N** | | | | |
| Non-Hispanic White | 4,932 | 27,428 | 1.0 | Reference |
| Non-Hispanic Black | 410 | 2,436 | 0.85 (0.76, 0.95) | 0.003 |
| Non-Hispanic Asian | 116 | 1,069 | 0.65 (0.54, 0.80) | <0.0001 |
| Non-Hispanic, Multi-races | 445 | 1,855 | 1.27 (1.14, 1.42) | <0.0001 |
| Hispanic | 780 | 4,142 | 0.97 (0.89, 1.05) | 0.467 |
| **Age, N** | | | | |
| 18-34 | 1,494 | 6,803 | 1.0 | Reference |
| 35-49 | 2,272 | 12,204 | 0.85 (0.80, 0.92) | <0.0001 |
| 50-64 | 1,668 | 11,414 | 0.65 (0.60, 0.71) | <0.0001 |
| 65 and above | 1,249 | 6,509 | 0.88 (0.81, 0.96) | 0.004 |
| **Education, N** | | | | |
| High School and Below | 1,153 | 5,480 | 1.0 | Reference |
| Some College/Associate | 3,037 | 13,905 | 1.03 (0.96, 1.11) | 0.41 |
| Bachelor's degree | 1,486 | 9,867 | 0.72 (0.66, 0.78) | <0.0001 |
| Graduate degree | 1,007 | 7,678 | 0.63 (0.58, 0.69) | <0.0001 |
| **Region, N** | | | | |
| Northeast | 800 | 4,968 | 1.0 | Reference |
| South | 2,389 | 11,990 | 1.21 (1.11, 1.32) | < 0.0001 |
| Midwest | 1,449 | 8,452 | 1.01 (0.92, 1.11) | 0.88 |
| West | 2,045 | 11,520 | 1.06 (0.97, 1.15) | 0.24 |

Odds ratios are calculated using adjusted logistic regression analyses that are controlled for the other categorical variables in the table.

**Table S3.** Prevalence of Cognitive Deficit Reported by Long COVID Patients in US States

| State | Cognitive Deficit | | % | State | Cognitive Deficit | | % |
|---|---|---|---|---|---|---|---|
| | Yes | No | | | Yes | No | |
| Alabama | 127 | 612 | 17.2 | Alaska | 98 | 546 | 15.2 |
| Arizona | 167 | 1033 | 13.9 | Arkansas | 142 | 629 | 18.4 |
| California | 349 | 2306 | 13.2 | Colorado | 182 | 964 | 15.9 |
| Connecticut | 77 | 584 | 11.6 | Delaware | 51 | 292 | 14.9 |
| District of Columbia | 53 | 291 | 15.4 | Florida | 187 | 1070 | 14.9 |
| Georgia | 159 | 875 | 15.4 | Hawaii | 46 | 314 | 12.8 |
| Idaho | 150 | 799 | 15.8 | Illinois | 124 | 803 | 13.4 |
| Indiana | 123 | 769 | 13.8 | Iowa | 140 | 668 | 17.3 |
| Kansas | 125 | 753 | 14.2 | Kentucky | 155 | 635 | 19.6 |
| Louisiana | 110 | 560 | 16.4 | Maine | 58 | 318 | 15.4 |
| Maryland | 93 | 655 | 12.4 | Massachusetts | 120 | 809 | 12.9 |
| Michigan | 170 | 1020 | 14.3 | Minnesota | 127 | 783 | 14.0 |
| Mississippi | 100 | 505 | 16.5 | Montana | 136 | 726 | 15.8 |
| Montana | 113 | 536 | 17.4 | Nebraska | 118 | 683 | 14.7 |
| Nevada | 98 | 594 | 14.2 | New Hampshire | 80 | 521 | 13.3 |
| New Jersey | 99 | 642 | 13.4 | New Mexico | 126 | 683 | 15.6 |
| New York | 115 | 658 | 14.9 | North Carolina | 114 | 600 | 16.0 |
| North Dakota | 77 | 423 | 15.4 | Ohio | 126 | 639 | 16.5 |
| Oklahoma | 192 | 805 | 19.3 | Oregon | 189 | 865 | 17.9 |
| Pennsylvania | 144 | 843 | 14.6 | Rhode Island | 52 | 306 | 14.5 |
| South Carolina | 126 | 638 | 16.5 | South Dakota | 84 | 510 | 14.1 |
| Tennessee | 161 | 789 | 16.9 | Texas | 340 | 1770 | 16.1 |
| Utah | 202 | 1114 | 15.3 | Vermont | 55 | 287 | 16.1 |
| Virginia | 151 | 752 | 16.7 | Washington | 218 | 1251 | 14.8 |
| West Virginia | 128 | 512 | 20.0 | Wisconsin | 99 | 675 | 12.8 |
| Wyoming | 109 | 515 | 17.5 | | | | |

Numbers in % indicate the percentages of long COVID patients reporting severe cognitive deficits in each US state.

**Table S4.** Sociodemographic characteristics of long COVID patients in CT, MD, KY and WV States

| | **CT**<br>**N = 661** | **MD**<br>**N= 748** | **KY**<br>**N = 790** | **WV**<br>**N = 640** | **p-value** |
|---|---|---|---|---|---|
| **Gender at Birth, N (%)** | | | | | 0.89 |
| Male | 193 (29.2) | 224 (29.9) | 224 (28.4) | 182 (28.4) | |
| Female | 468 (70.8) | 524 (70.1) | 566 (71.6) | 458 (71.6) | |
| **Race/Ethnicity, N (%)** | | | | | < 0.0001 |
| Non-Hispanic White | 467 (70.7) | 446 (59.6) | 708 (89.6) | 592 (92.5) | |
| Non-Hispanic Black | 49 (7.4) | 164 (21.9) | 26 (3.3) | 13 (2.0) | |
| Non-Hispanic Asian | 15 (2.3) | 27 (3.6) | 8 (1.0) | 6 (0.9) | |
| Non-Hispanic, Multi-races | 22 (3.3) | 32 (4.3) | 25 (3.2) | 13 (2.0) | |
| Hispanic | 108 (16.3) | 79 (10.6) | 23 (2.9) | 16 (2.5) | |
| **Age, N (%)** | | | | | 0.23 |
| 18-34 | 123 (18.6) | 122 (16.3) | 154 (19.5) | 95 (14.8) | |
| 35-49 | 230 (34.8) | 257 (34.4) | 246 (31.1) | 221 (34.5) | |
| 50-64 | 209 (31.6) | 246 (32.9) | 246 (31.1) | 223 (34.8) | |
| 65 and above | 99 (15.0) | 123 (16.4) | 144 (18.2) | 101 (15.8) | |
| **Education, N (%)** | | | | | < 0.0001 |
| High School and Below | 112 (16.9) | 102 (13.6) | 147 (18.6) | 154 (24.1) | |
| Some College/Associate | 229 (34.6) | 213 (28.5) | 326 (41.3) | 254 (39.7) | |
| Bachelor's degree | 175 (26.5) | 202 (27.0) | 176 (22.3) | 129 (20.2) | |
| Graduate degree | 145 (21.9) | 231 (30.9) | 141 (17.8) | 103 (16.1) | |

N represents the total number of long COVID patients in each of the four states. The numbers inside the parenthesis indicate the percentages of long COVID patients in each sociodemographic group. P-value calculation was based on the Chi-Squared test.
